# Using genomic epidemiology and geographic activity spaces to investigate tuberculosis outbreaks in Botswana

**DOI:** 10.1101/2024.12.04.24318520

**Authors:** Chelsea R. Baker, Ivan Barilar, Leonardo S. de Araujo, Daniel M. Parker, Kimberly Fornace, Patrick K. Moonan, John E. Oeltmann, James L. Tobias, Volodymyr M. Minin, Chawangwa Modongo, Nicola M. Zetola, Stefan Niemann, Sanghyuk S. Shin

## Abstract

**Background:** The integration of genomic and geospatial data into infectious disease transmission analyses typically includes residential locations and excludes other activity spaces where transmission may occur (e.g. work, school, or social venues). The objective of this analysis was to explore residential as well as other activity spaces of tuberculosis (TB) outbreaks to identify potential geospatial ‘hotspots’ of transmission.

**Methods:** We analyzed data that included geospatial coordinates for residence and other activity spaces collected during 2012–2016 for the Kopanyo Study, a population-based study of TB transmission in Botswana. We included participants with results from whole genome sequencing conducted on archived samples from the original study. We used a spatial log-Gaussian Cox process model to detect core areas of increased activity spaces of individuals belonging to TB outbreaks (genotypic groups with ≤5 single-nucleotide polymorphisms), which we compared to ungrouped participants (those not in a genotypic group of any size).

**Findings:** We analyzed data collected from 636 participants, including 70 participants belonging to six outbreak groups with a combined total of 293 locations, and 566 ungrouped participants with a combined total of 2289 locations. Core areas of activity space for each outbreak group were geographically distinct, and we found evidence of localized transmission in four of six outbreaks. For most of the outbreaks, including activity space data led to the detection of larger areas of higher spatial intensity and more focal points compared to residential location alone.

**Interpretation:** Geospatial analysis using activity space data (social gathering places as well as residence) may lead to improved understanding of areas of infectious disease transmission compared to using residential data alone.

**Funding:** This work was supported by funding from the National Institute of Allergy and Infectious Diseases R01AI097045, R01AI147336, and R01AI170204.

## Background

Tuberculosis (TB) remains among the leading causes of death due to infectious illness, despite being a preventable and curable disease^1^. In 2022, over 10 million people became sick with TB, and 1.3 million died^1^. Progress toward TB elimination has been slow and many targets set by the World Health Organization (WHO) have not been met^1^. New strategies and tools are needed for TB prevention^1^. In high-burden settings, where a substantial portion of disease incidence is due to recent infection, interventions to stop ongoing transmission are especially important^1–5^.

A promising tool is the integration of geospatial and pathogen genomic data. Pathogen whole genome sequencing (WGS) can be used to identify closely related *M. tuberculosis* isolates and help reconstruct likely transmission chains^6^. Geographic and genomic data can be combined to help detect areas of sustained transmission, locate outbreaks, and investigate the geographic range of different strains^7,8^. Spatial analysis of WGS data can help identify high-risk areas that could be targeted for public health interventions to interrupt ongoing transmission^3–7,9–15^.

Geographically targeted interventions have shown promise as an effective and cost-efficient strategy for reducing TB incidence in high-burden, low-resource settings^16–19^.

However, an important limitation to many studies employing this strategy is geospatial analysis based solely on residential location, which excludes locations in the community where transmission may occur^12,20–22^. An alternative approach is to analyze “activity space,” which includes the places one routinely occupies during day to day life^23–25^. For example, this may include residential as well as community sites such as workplaces, markets, places of worship, or other social gathering places^23,24^. This approach has the potential to lead to more accurate detection of high-risk areas compared to analysis of residential locations alone^24,26^.

We previously conducted a descriptive study of geospatial residential data and WGS data from a population-based study of TB transmission in Botswana found evidence that TB outbreaks displayed distinct geographic characteristics^27^. The objectives of the current analysis were to use spatial statistical modeling to 1) identify geographic characteristics of the collective activity space (residential as well as social gathering locations) of each outbreak group, and 2) identify potential ‘hotspot’ areas of activity space associated with each outbreak, which may represent areas of increased risk for transmission.

## Methods

### Study design and setting

We analyzed data collected during 2012–2016 for the Kopanyo Study, a population-based study of TB transmission in Botswana, a country in southern Africa with a high burden of TB and TB/HIV co-infection^1,5,28^. Participants were recruited at multiple local health clinics in two districts: Gaborone, the urban center and capital city, and Ghanzi, a rural district several hundred kilometers away^5,28^. During the five years before the study, TB incidence was 440–470 cases/100,000 persons in Gaborone, which had a total population of 354,380, and 722 cases/100,000 persons in Ghanzi, which had a population of 44,100 (12,179 in Ghanzi town)^5,28^.

Study participants included men and women of all ages with TB disease who were sequentially enrolled by date of diagnosis^5,28^. Those who had already received TB treatment for >14 days, prisoners, and patients who declined to participate were excluded^5,28^. At least 1 sputum sample was collected from each participant for bacterial culture^5,28^. Clinical and demographic data were collected through in-person interviews and medical record review^5,28^.

Data gathered during participant intake interviews included high resolution geospatial data for activity space, which included home residence and social gathering places^5,28^. Participants were asked about residential location as well as social gathering places (e.g. workplaces, schools, markets, places of worship, alcohol venues etc.) frequented during their potential infectious period (up to 12 months prior to treatment initiation)^5,28^. Geographic coordinates (latitude and longitude recorded using the WGS 84 projection system with 1.1-m precision) for locations were obtained using global positioning system (GPS) devices during site visits, or by geocoding addresses using a reference layer created by manually relocating addresses in satellite imagery using Google Maps, OpenStreetMap, and ArcGIS^5,28^.

### WGS

Whole genome sequencing was conducted on DNA samples archived from the original study with sufficient quantities of DNA (>0.05 ng/μL) for analysis. Closely related *M. tuberculosis* isolates were identified bioinformatically using a single linkage clustering algorithm. We considered clusters of isolates with ≤5 single-nucleotide polymorphisms (SNPs) to indicate recent transmission and clusters of ≥10 persons to be outbreaks. Further details of this procedure are outlined in a separate analysis^27^.

### Spatial modeling of activity space

Participants eligible for the current analysis included those with WGS data, GPS coordinates, and sociodemographic data for age, sex, income, and HIV status available. We focused our current analysis on outbreak groups that had at least 10 activity space locations (collectively among all their participants) within greater Gaborone, an area of approximately 27 km x 24 km that includes the capital city and its surrounding suburbs. We also included genotypically ungrouped participants as a comparison group. For model fitting purposes, a very small jitter was introduced to location coordinates to avoid duplicate points (roughly on the scale of different areas of the same building, ranging from approximately <1 to 10 meters).

We conducted a preliminary analysis to compare the geographic distribution of participants with WGS data available to the total study population from the Kopanyo Study to rule out geographic sampling bias. We estimated the geographic median center (a centralized point that minimizes the distance to all other points), and directional distribution (which calculates the standard deviation of points along both the X and Y axes) for both groups of participants and found nearly identical results, indicating that participants with WGS data were geographically representative of the larger study population. This analysis was performed using ArcGIS^29^.

### Model description

We used a spatial log-Gaussian Cox process (LGCP) to model the spatial intensity (average number of points per unit area) of activity spaces of participants belonging to each outbreak group (‘cases’) and of genotypically ungrouped participants (those not in an identified genotypic group of any size, ‘controls’). LGCPs are a flexible class of models for spatial point processes where spatial intensity may vary across the study region^30,31^. A spatial random effect can be incorporated to account for spatial correlation in the data and identify spatial patterns not explained by other variables^33,34,37,40^. This technique offers a model-based approach for estimating utilization distributions, which are probability density functions that can be mapped to highlight areas with increased geographic concentrations of points (e.g. activity space locations) to help characterize use of space^26,32^. To adapt the modeling framework to an activity space context where each individual may be associated with multiple point locations, observations can be treated as cumulative ‘encounters’ over specified time periods^32^. We considered each point to represent an ‘encounter’ in space corresponding to a potential TB exposure, and estimated intensity surfaces for cumulative exposures over the entire study period.

LGCPs fit well in a Bayesian hierarchical modeling framework, and various tools can be used for this approach^33–35^. We used integrated nested Laplace approximation (INLA), a flexible and computationally efficient method for approximate Bayesian inference for latent Gaussian models, which include LGCPs^33,34,36–38^. We implemented this using the R-INLA package^39^. We modeled the spatial random effect as a Gaussian random field (GRF) with Matérn covariance^37,38,40^. We used the stochastic partial differential equation (SPDE) approach in R-INLA to approximate the GRF^37,38,40^. We specified the SPDE model using penalized complexity priors that were vaguely informative about the underlying spatial process (prior probability of 0.05 that the ranges of the fields were less than 0.5 km and prior probability of 0.05 that the standard deviation was greater than 10).

Under the LGCP framework, we used a joint modeling approach to incorporate a shared spatial term (obtained by jointly estimating the intensity of both cases and controls), as well as a unique spatial term estimated for cases in each group^38,41^. Using this approach, posterior mean estimates of the spatial random effect for cases represent variation in intensity not accounted for by the spatial distribution of controls^41^. We did this to help identify areas with relatively high concentrations of activity spaces associated with individual outbreak groups, while attempting to account for baseline use of space (as some locations tend to be frequented by people more often in general). Areas with a high density of activity spaces frequented by people belonging to the same outbreak group could potentially represent areas associated with an increased risk of recent transmission. We fit a version of the model that included just the shared spatial term (model 0), and a version of the model that included the shared spatial term as well as unique spatial terms estimated for each outbreak group individually (model 1). We also conducted a sensitivity analysis using subsets of the data with 70 and 140 randomly selected ungrouped participants as controls to examine whether spatial patterns were sensitive to size of the control group.

We projected posterior mean estimates of the spatial effect (i.e. the effect of spatial location on the intensity of activity spaces, represented by the spatial random field) for each outbreak group onto maps of the study area in order to visualize how it varied across the region, and to identify areas of increased or decreased (different than zero) values not explained by the spatial distribution of controls^41^. Estimated values (displayed on the internal linear predictor scale) represent the contribution of the spatial random effect to the response (spatial intensity) after accounting for other fixed and random effects in the model. In addition, we reported posterior mean estimates for the range (distance at which spatial correlation falls close to zero) and variance of the spatial effect for each outbreak group^36^.

We also projected posterior mean estimates for predicted spatial intensity values (fitted values of the response at prediction locations, obtained by exponentiating the linear predictor), in order to visualize patterns of spatial intensity of activity spaces for each outbreak group^36,38^.

In addition, we calculated exceedance probabilities and projected these onto maps of the study area to identify areas where estimated spatial effect for each outbreak group had a high probability (0.95) of being greater than zero, representing high-confidence areas where the spatial effect for cases was above the baseline that could be accounted for by the spatial distribution of controls^41^. We also calculated exceedance probabilities to identify high-confidence areas where the estimated spatial intensity was in the top ten percent of estimated mean values for each group, representing ’core areas’ or ‘hotspots’ of that group’s collective activity space^32^.

We also generated exceedance probability maps based only on location of participant residence (using the same threshold values as the full analysis) to compare high-risk areas identified using activity space analysis vs. home location alone. We projected these exceedance probabilities onto interactive maps of the study area for each outbreak group.

Map visualization was performed using the R packages raster, terra, sf, ggplot2, ggspatial, and leaflet.

## Results

### Participants

A total of 1426 participants had WGS data available, of which 1425 had GPS coordinates available for at least one activity space location (home or social gathering place). Participants with and without WGS data had similar sociodemographic characteristics in terms of age, sex, HIV status, and income. Eight genotypic groups had 10 participants or more and were considered outbreaks. Six out of eight outbreaks (genotypic groups with 10 participants or more) had at least 10 activity spaces (collectively among all their participants) in greater Gaborone.

A total of 636 participants with activity spaces in greater Gaborone met criteria for the current analysis, including 70 participants belonging to six outbreak groups with a combined total of 293 locations, and 566 ungrouped participants with a combined total of 2289 locations.

Each participant had between one and 10 activity space locations, and the median number of locations (n=4) was the same for both grouped and ungrouped participants (Figure 1). Median number of activity space locations was the same (n=4) by gender and HIV status, though was slightly lower for participants with no income (n=3) than participants with any income (n=4), which could reflect an increased number of activity spaces among participants who were employed. Among participants with more than one location, the maximum distance between any two of their activity spaces ranged from <0.5 km to 21.2 km (median 6.2 km) for ungrouped participants and <0.5 km to 21.7 km (median 4.3 km) for participants in outbreak groups (supplementary figure 1).

**Figure 1.**
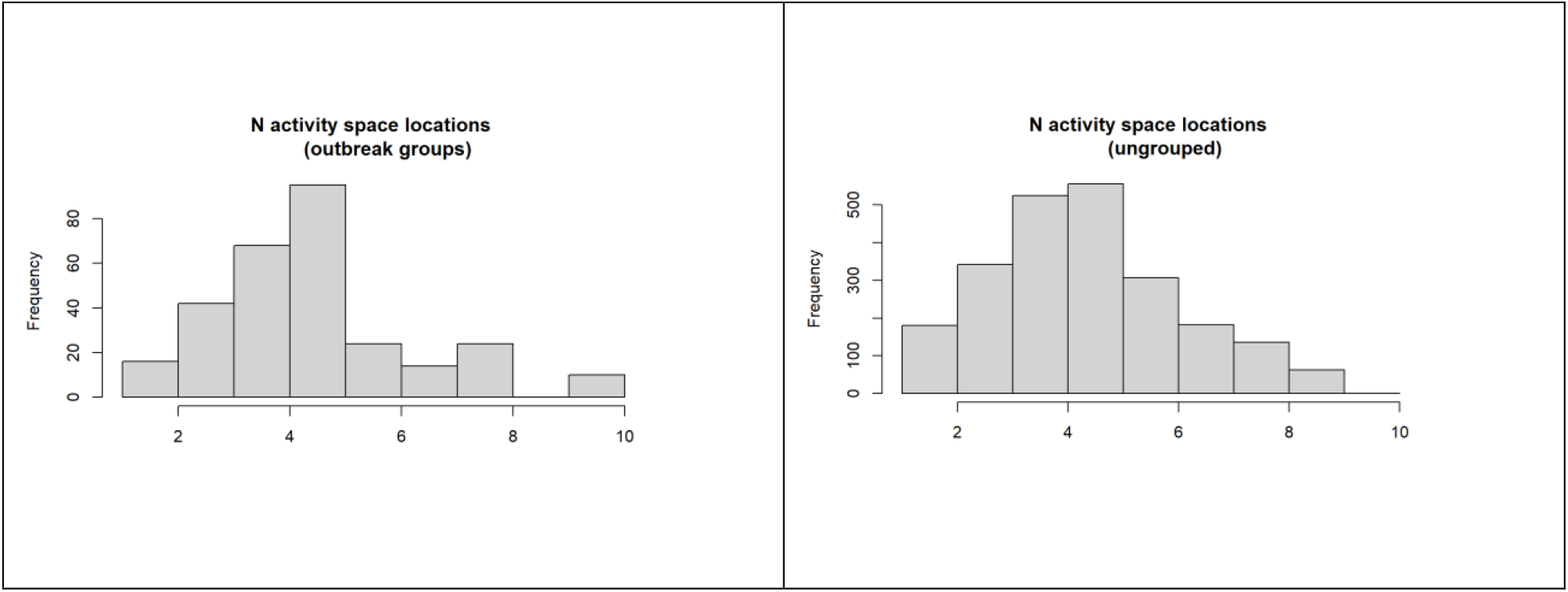
Histograms for distribution of number of activity space locations per participants for outbreak and genotypically ungrouped participants.

Among genotypically ungrouped participants, the median age was 35 years (IQR: 28–42), just over half were male, about one quarter reported no income, and nearly 65% were diagnosed with TB-HIV coinfection (Table 1). Among participants in the six genotypic groups, median age ranged from 30 years (Group A) to 39 years (Group G) (Table 1). Participants in Group G were exclusively male, while Group C alone was majority female (75%). Group D had the highest proportion of participants diagnosed with TB-HIV coinfection (9 of 11; 91%). The percentage of participants reporting no income ranged from 18% in Group D to 58% in Groups C and E (Table 1).

**Table 1.**
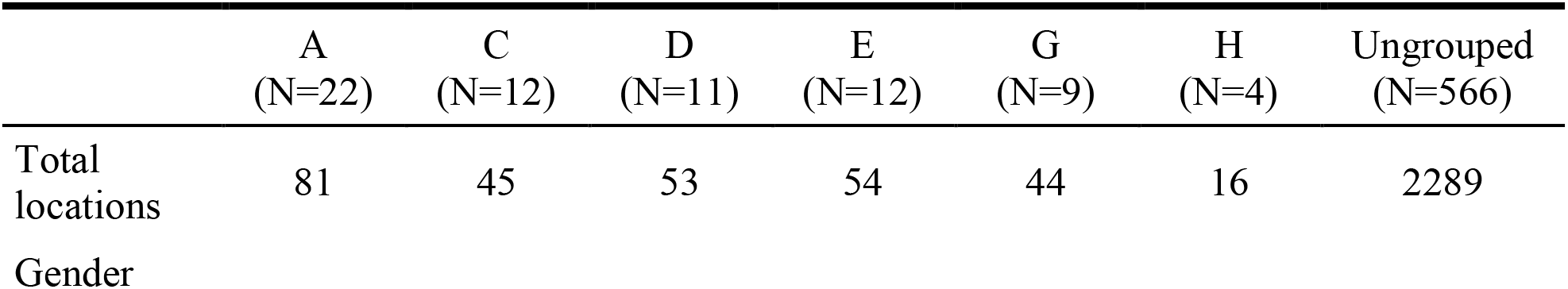
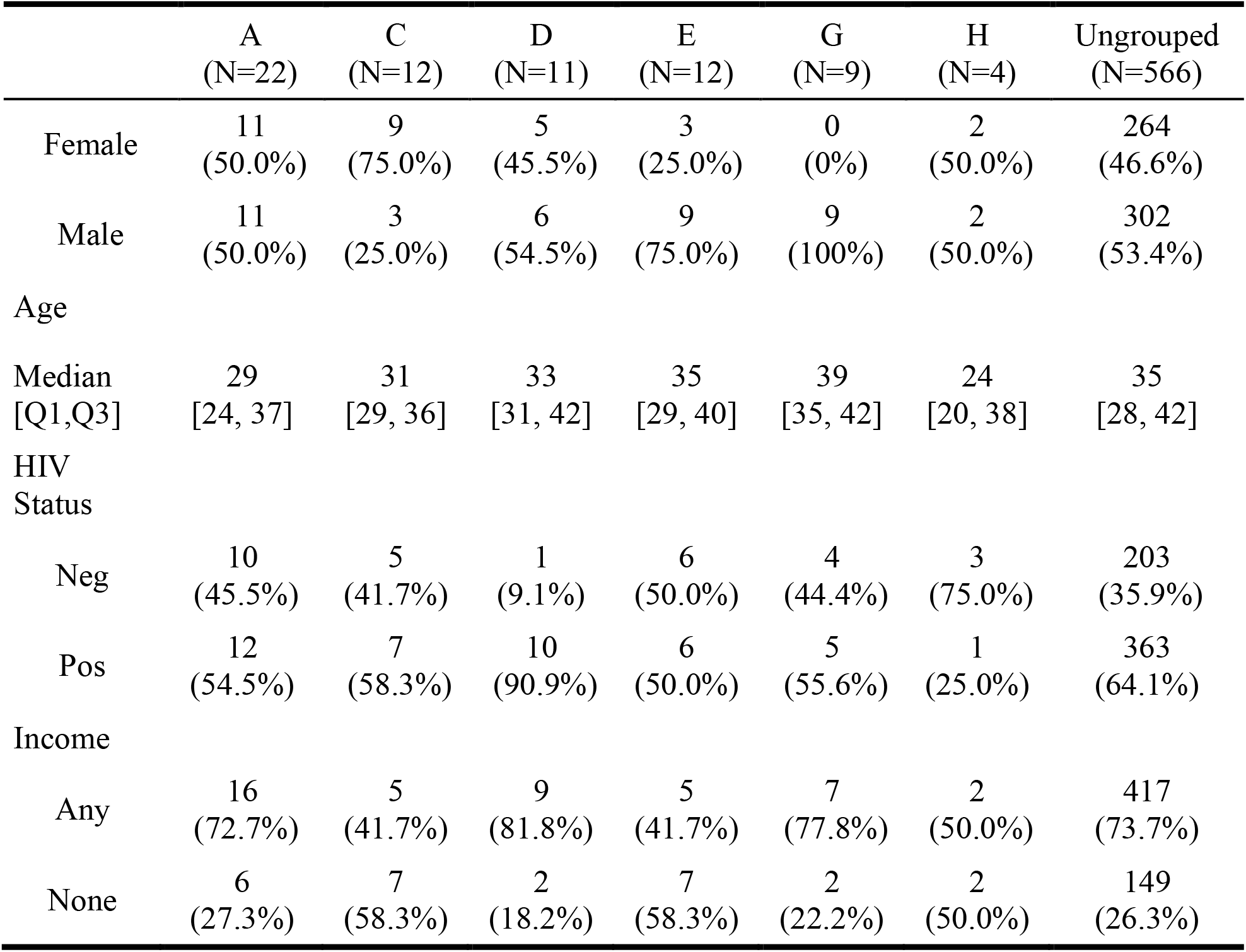
Characteristics of study participants (N = 636) by outbreak group (genotypic group ≤ 5 SNP), Gaborone, Botswana, 2012-2016

### Estimated spatial effects

Model 1 (shared and group-specific spatial terms) had a lower DIC (-12956.58) than model 0 (shared spatial terms only, DIC -12853.84), supporting the presence of spatial variation among genotypic groups not accounted for by the spatial distribution of activity spaces of controls^41^.

In general, posterior estimates for the range of the spatial effects suggested small to medium scale spatial correlation (Table 2; Figure 2). The range was smallest for groups A and H, indicating that spatial correlation among points died off at relatively short distances. Both the range and variance were largest for group C, indicating the spatial effect spanned a greater distance but also displayed ‘peaks’. This could be due to the presence of two distinct areas of increased intensity located relatively far from one another.

**Figure 2.**
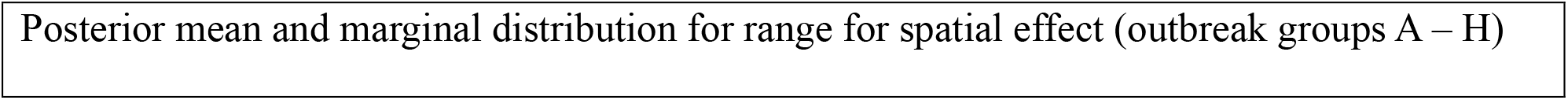
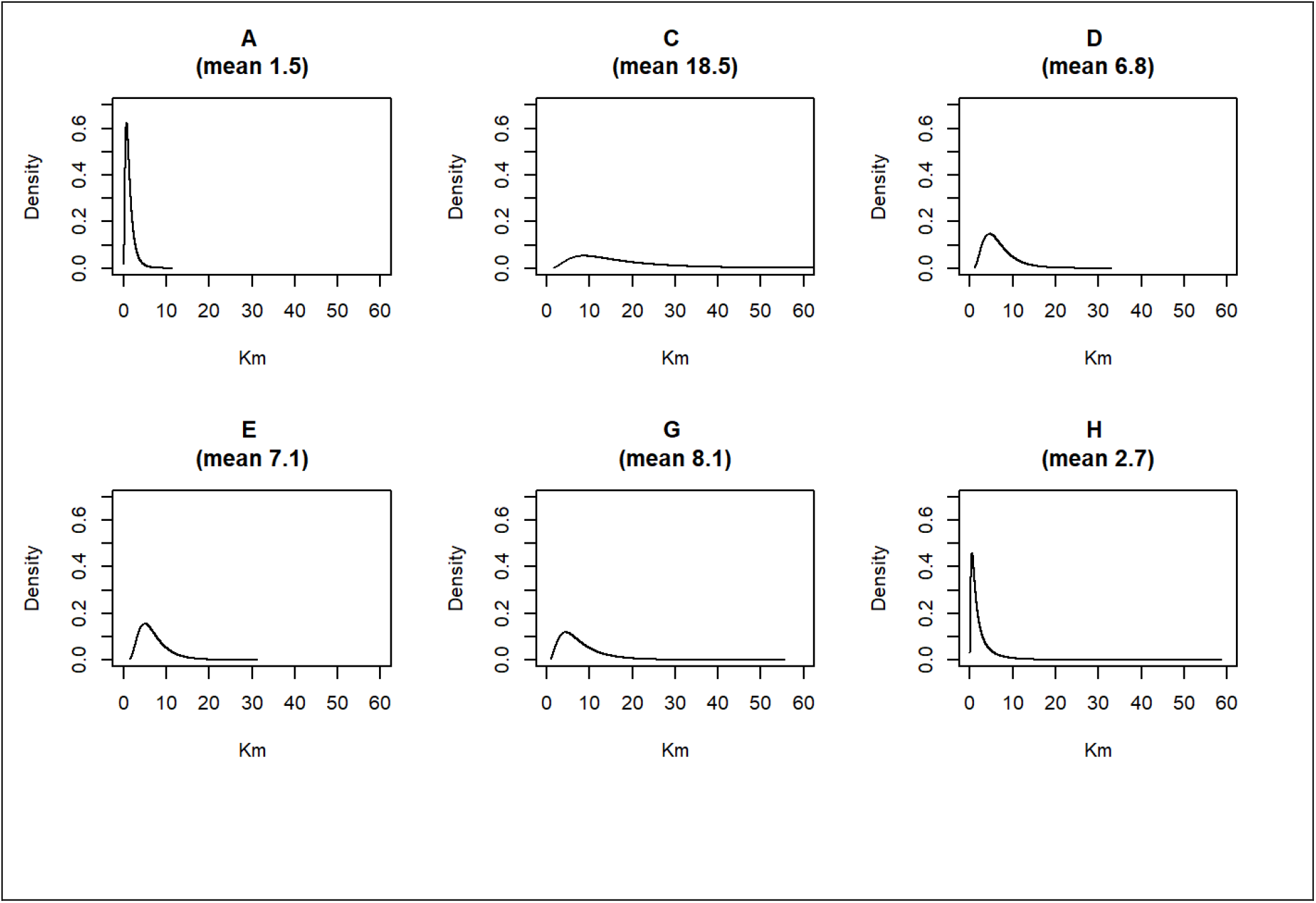
Posterior mean estimates and marginal distributions of the range and variance of the estimated spatial effect for each outbreak group, Gaborone, Botswana, 2012-2016

**Table 2.**
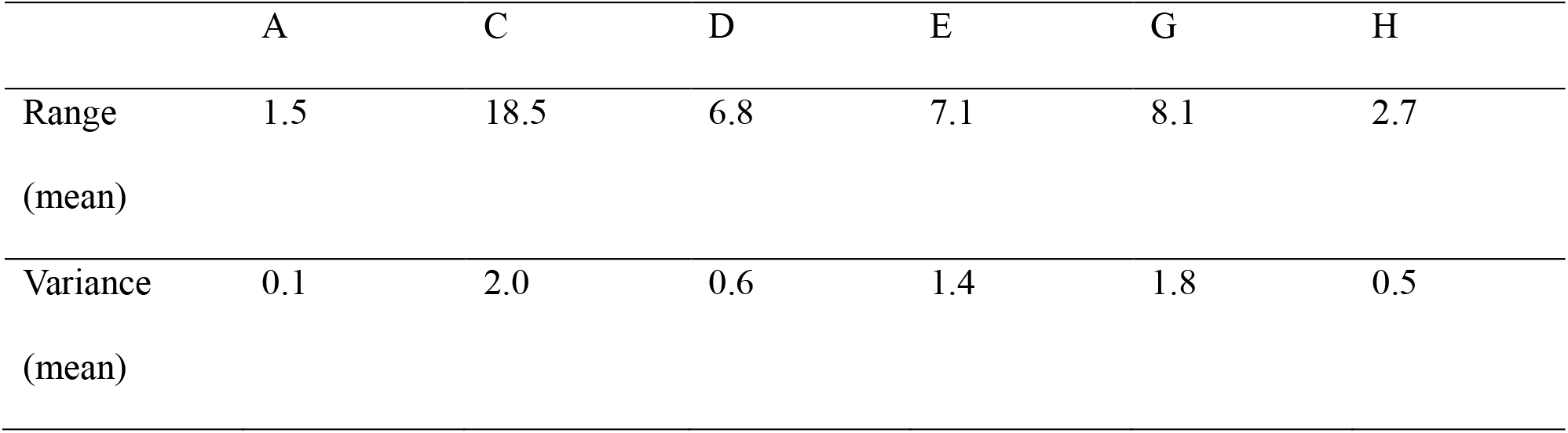
Posterior mean estimates of the range and variance of the spatial effect for each outbreak group (A-H), Gaborone, Botswana, 2012-2016.

Maps of posterior mean estimates of spatial effects displayed different spatial patterns for each outbreak group (Figure 3). Estimated spatial effects for group A and group H showed several relatively small and dispersed areas of increased values compared to controls. Group C had a notable area of increased spatial effect in the central southern part of the study area. Group D and group G had two to three main areas of increased values that followed a broad east-west spread, while for group E areas of increased estimates had a general north-south configuration.

**Figure 3.**
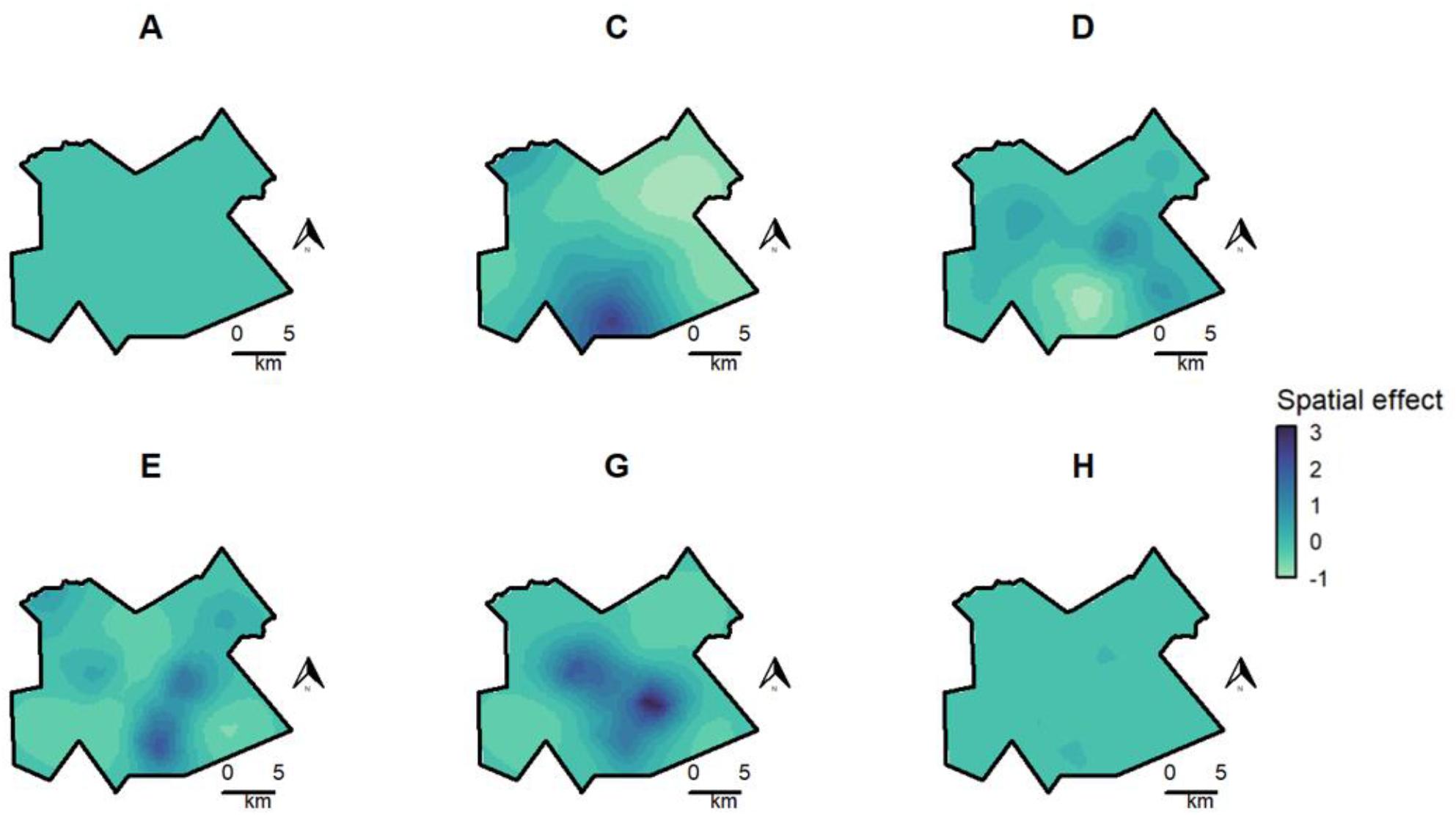
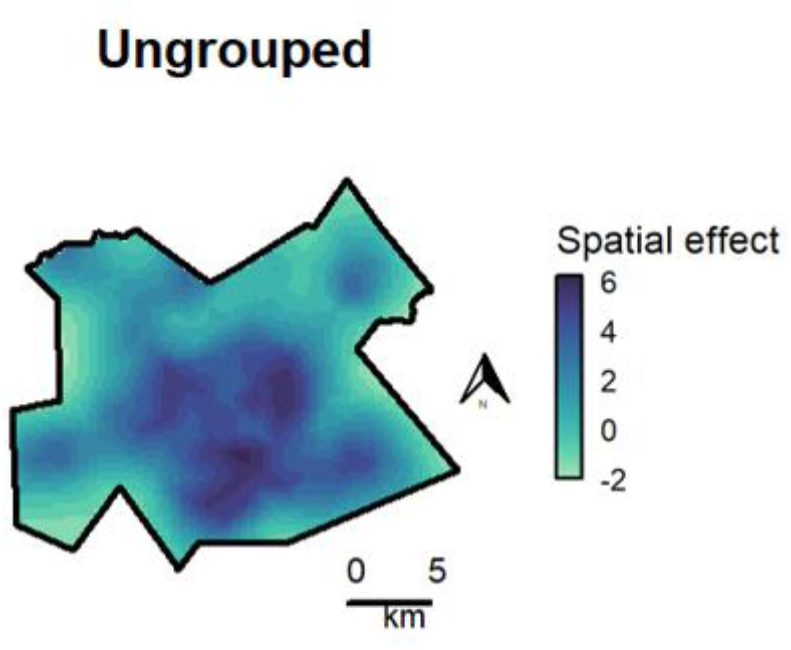
Posterior mean estimates of spatial random effect for each outbreak group (A-H) and controls (ungrouped participants), Gaborone, Botswana, 2012-2016. Values are shown on the internal linear predictor scale and represent the contribution of the spatial random effect on the response, after accounting for other fixed and random effects in the model. Departures from baseline (above or below zero) for outbreak groups measure group-specific spatial patterns that are not accounted for by the spatial distribution of activity spaces of controls. Darker colors correspond to increased spatial effect estimates. Values are displayed on the same color scale for all outbreak groups, though on a separate color scale for controls due to difference in sample size.

Results of the sensitivity analysis using subsets of 70 and 140 randomly selected controls found very similar results in terms of the spatial patterns and magnitude of estimated spatial effect by group (Supplementary Figure 2 and Supplementary Figure 4).

### Predicted spatial intensity

Maps of predicted mean spatial intensity displayed unique spatial patterns for each outbreak group (Figure 4).

**Figure 4.**
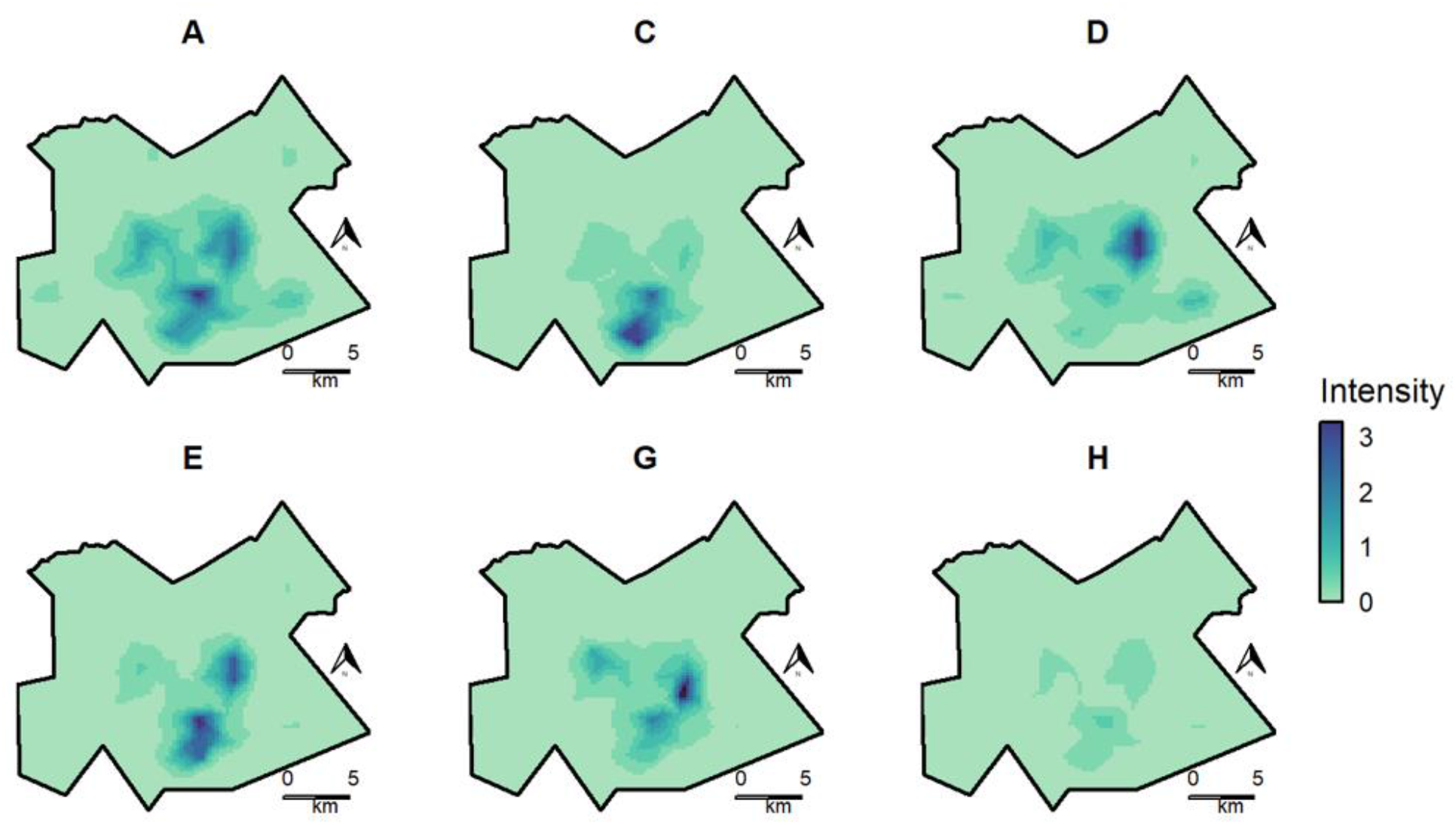
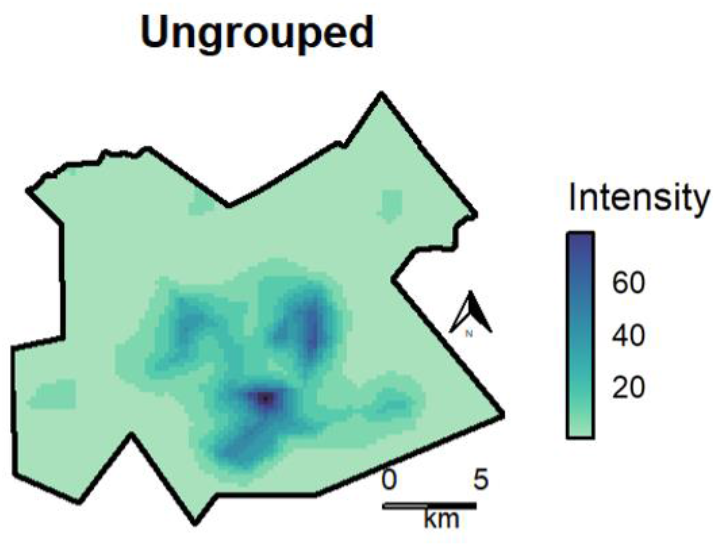
Predicted mean spatial intensity of activity spaces for participants in each outbreak group (A-H) and controls (ungrouped participants), Gaborone, Botswana, 2012-2016. Values are displayed on the response scale (obtained by exponentiating the linear predictor) and represent predicted numbers of activity spaces per unit area (approximately 0.25 x 0.25 km). Areas of increased intensity correspond to higher geographic concentration of activity spaces for participants in each group. Intensity values are displayed on the same color scale for all outbreak groups for ease of visual comparison, though on a separate color scale for controls due to difference in sample size.

For group A, areas of increased spatial intensity of activity spaces followed an overall similar pattern as seen for controls, with areas of highest intensity toward the center of the study area. Group C had a distinct area of high intensity in the central southern part of the study area. Group D had a notable area of increased intensity in the central east part of the map. Areas of highest intensity for group E were in the central and south east, and for group G in the central east. The areas of highest intensity for group H were located toward the center of the study area and also resembled the overall spatial pattern seen for controls, though predicted values were relatively small compared to the other groups and not easily visible when mapped on the same color scale.

Results of the sensitivity analysis using subsets of 70 and 140 randomly selected controls found very similar results for predicted spatial intensity by outbreak group (Supplementary Figure 3 and Supplementary Figure 5).

### Exceedance maps

Exceedance maps for estimated spatial effects showed areas where the posterior mean had a high probability (0.95) of being above 0 (greater than baseline) (Figure 5). Areas of significantly increased spatial effect estimates based on full activity space analysis were detected for groups C, D, E, and G. Groups A and H did not have areas meeting the specified threshold, which may be due to a spatial distribution of activity spaces that resembles that of the control group.

**Figure 5:**
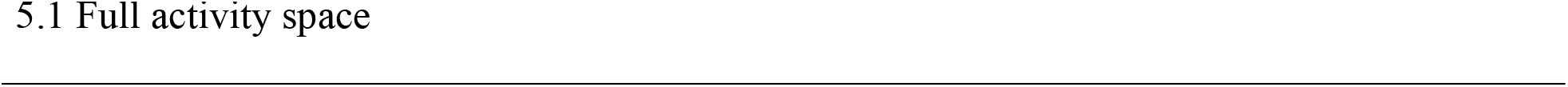
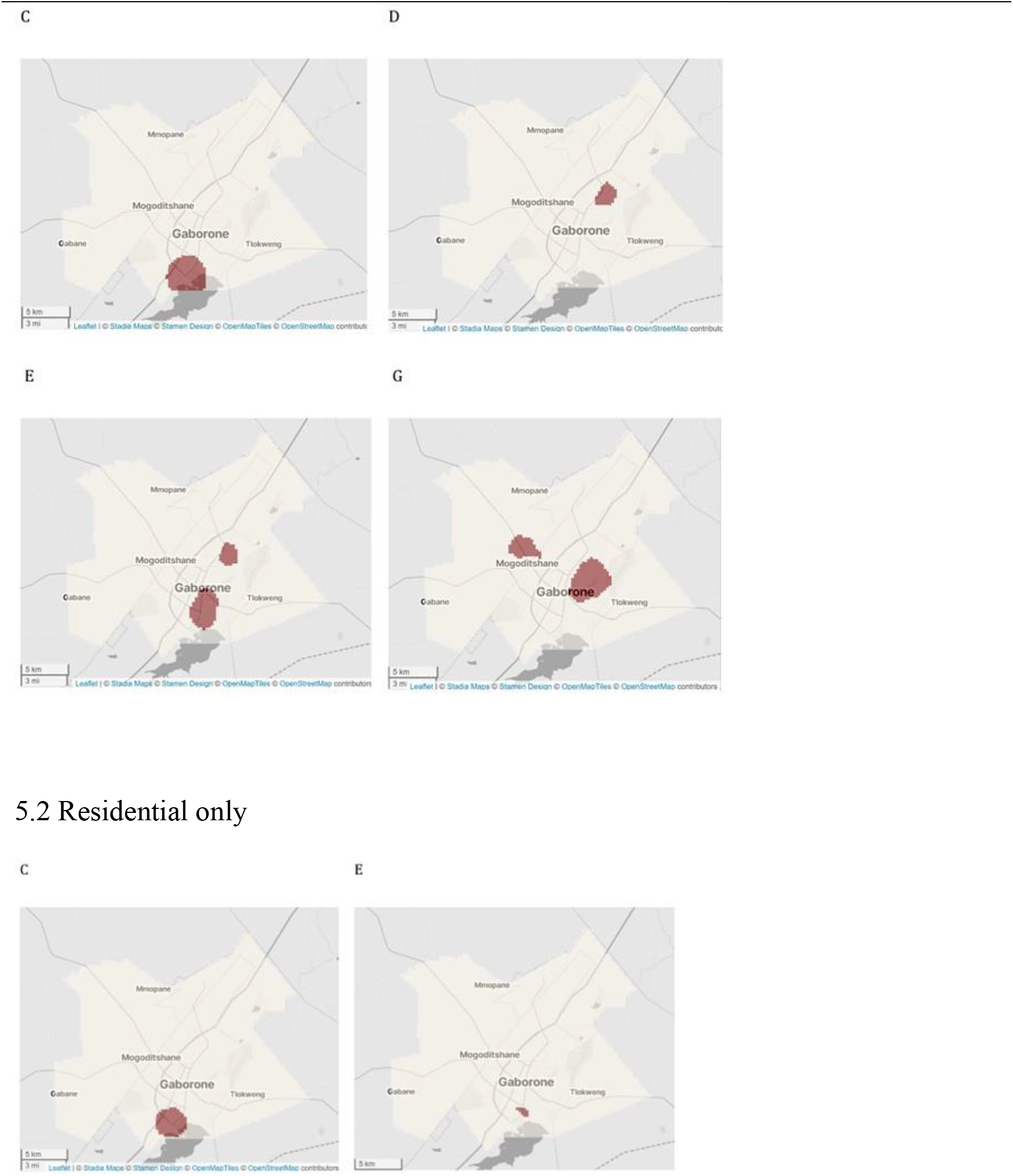
Exceedance maps for spatial effect greater than 0 (departure above baseline) with high probability (0.95) – full activity space and residential locations only, Gaborone, Botswana, 2012- 2016.

Areas of significantly increased spatial effect estimates based on residential location alone were detected for groups Cand E, though not for groups A, D, G, or H. For group E, exceedance areas based on activity space were geographically broader than those based solely on residential location. For group C, exceedance areas were similar in both analyses.

Exceedance maps for predicted spatial intensity values showed distinct areas of high spatial intensity (‘core areas’) for each outbreak group, corresponding to areas where posterior mean intensity values had a high probability (0.95) of being in the upper ten percent of estimates for that group (Figure 6).

**Figure 6.**
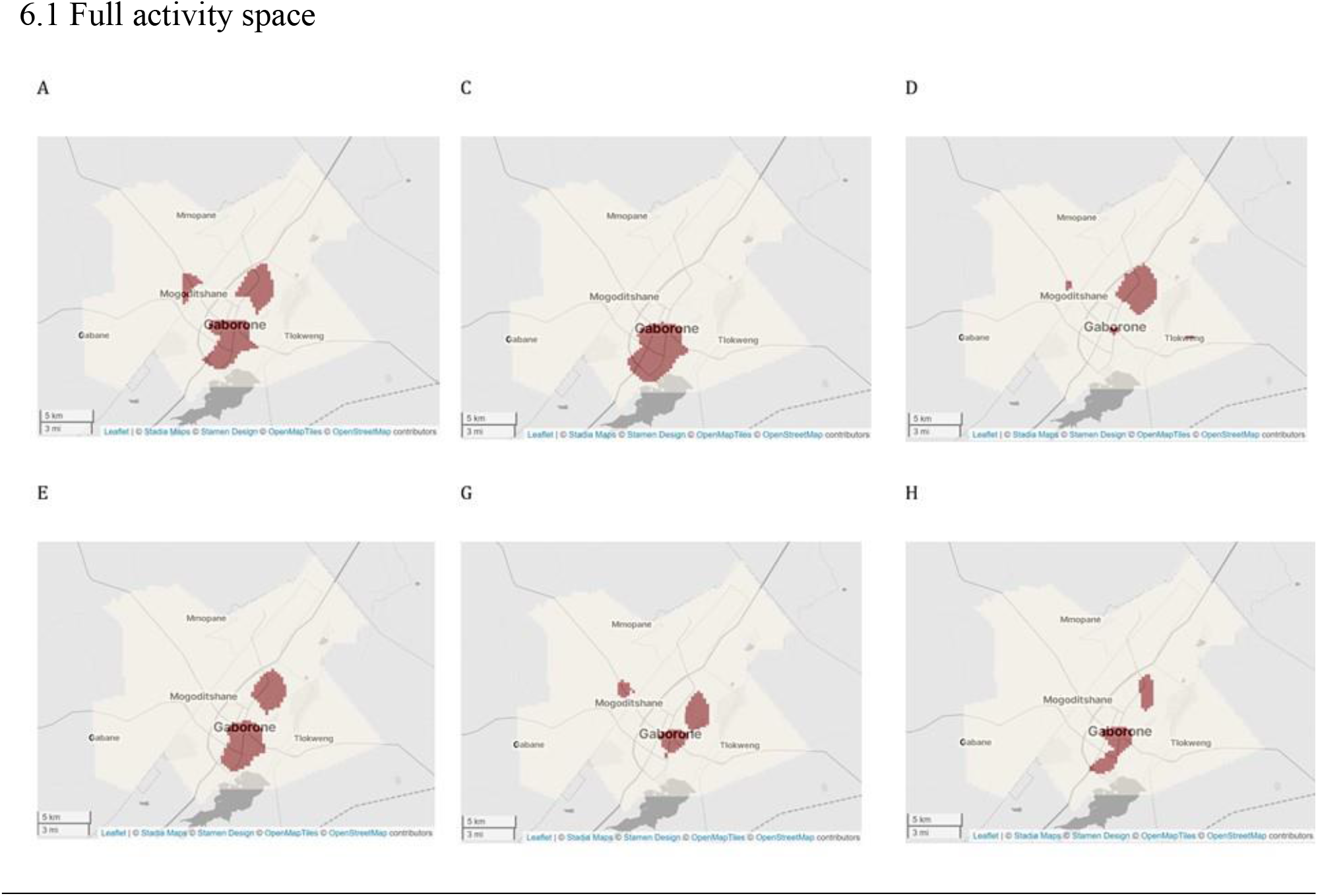
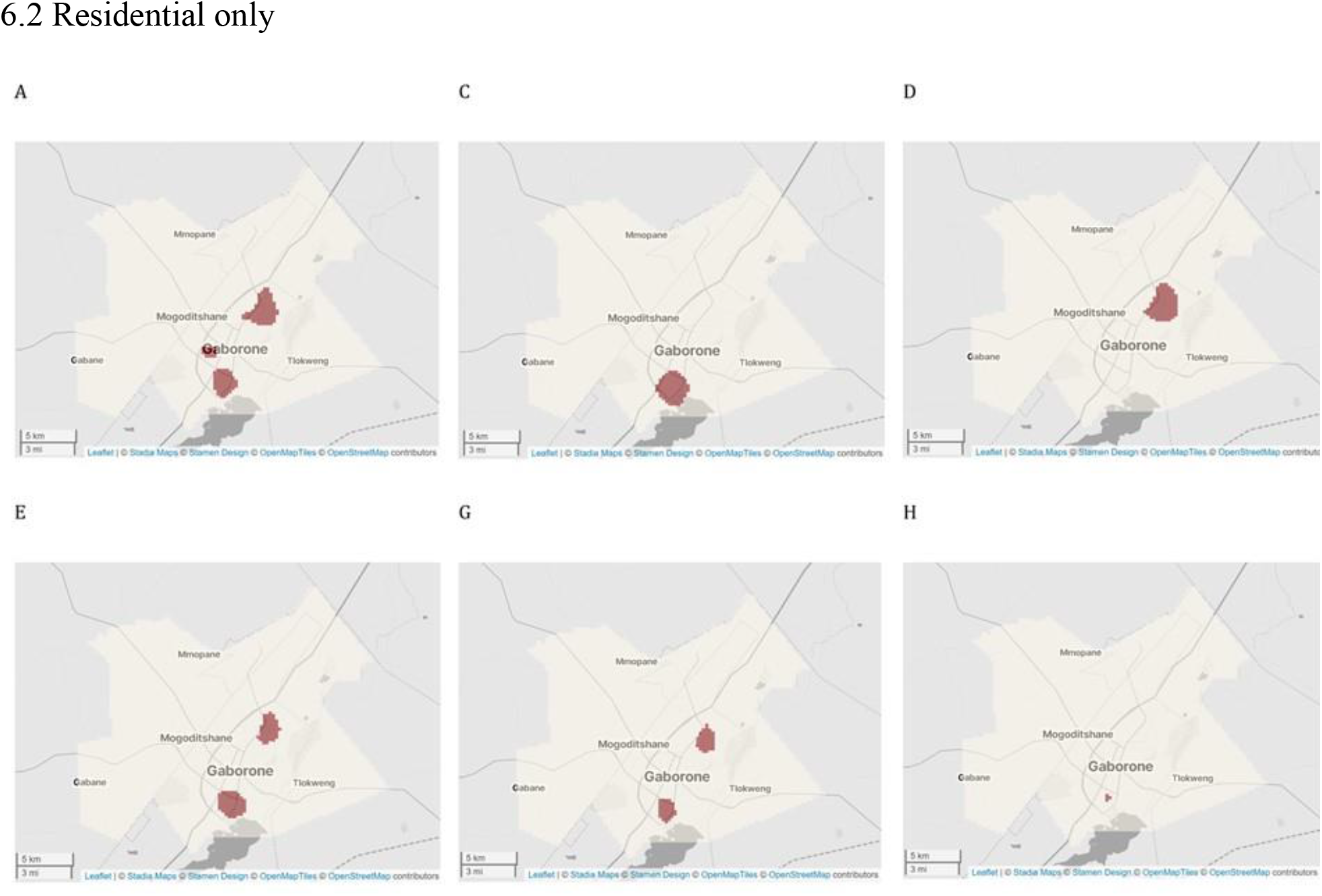
Exceedance maps for predicted spatial intensity to display ‘core areas’ by outbreak group based on full activity space and residential locations only, Gaborone, Botswana, 2012- 2016

In general, core areas based on residential location alone were geographically restricted compared to core areas based on full activity space analysis. For all groups core areas based on full activity space analysis were larger than those based on residential locations alone. For groups A, D, G, and H, core areas based on activity space also involved additional geographic focal areas.

## Discussion

In our analysis, we detected geographically distinct patterns of activity space associated with different TB outbreak groups. Core areas (‘hotspots’) of highest spatial concentration of activity spaces for each group were located in different areas, with some being more geographically widespread and others more compact. For outbreak groups C, D, E, and G, we detected areas where the spatial concentration of activity spaces of grouped participants was significantly higher than the baseline spatial distribution of activity spaces of ungrouped controls (increased spatial effect). This could suggest that distinct areas of localized transmission play an important role in these outbreaks. The spatial distribution of activity spaces for groups A (the largest outbreak group) and H (the smallest) resembled the overall spatial distribution of activity spaces belonging to the control group. The differences in spatial characteristics among the groups could potentially correspond to the timing of how long a genotype of TB has been circulating in the community. It could also represent transmission among socially or geographically distinct contact networks.

Activity space hotspots could represent potential high-priority areas for spatially targeted interventions such as active case finding for TB and other infectious diseases. This may be particularly useful for outbreaks involving localized transmission. Exceedance maps displaying core areas of spatial intensity and areas of increased spatial effects such as those shown above could potentially be a useful tool for public health planning. We displayed static snapshots from interactive maps at a relatively low spatial resolution to protect privacy, though in practice such maps could be used to examine potential hotspots at different spatial scales.

We also found differences between exceedance areas detected using full activity space analysis compared to residential location alone. Areas of core spatial intensity and significant spatial effects based on activity space were generally larger, and sometimes included additional geographic focal points, suggesting a notable portion of activity spaces may be located in areas outside participants’ home neighborhoods. A possible exception is group C, which had similar exceedance areas in both analyses, suggesting activity spaces for these participants may generally be located in closer proximity to place of residence. Relatively high unemployment in group C and fewer work locations may have contributed to this observation. Our results show that analyses based on residential location alone may not fully represent the spatial characterization of hotspots.

Spatial analysis for infectious disease transmission involves an inherent assumption that the locations analyzed are important with regard to transmission. Activity space analysis incorporates important locations in the community where TB transmission may occur, and may reduce exposure misclassification and improve the geographic characterization of transmission chains^42^. This has implications for planning and evaluating targeted interventions^43^. For example, a recent TB modeling study found limited effectiveness of spatially targeted screening based on proximity to household locations of people with incident TB in Peru^44^. However, hotspots of TB incidence based on household locations do not necessarily correspond with hotspots of transmission^43,44^. Activity space analysis may help address issues such as this in spatial analysis for TB transmission.

Activity space analysis has an established history of use in fields such as social geography and urban planning^45–47^. It fits naturally into the spatial epidemiology framework, which emphasizes place and location-based health exposures^23,48^. This approach acknowledges space as a social determinant of health and helps incorporate the influence of social and physical environments on health outcomes^23,49^. However, there are relatively few examples of spatial analysis of activity space in the TB literature. An early example that helped lay the foundation for activity space analysis in TB research was a study to detect TB hotspots in Japan^50^. The study incorporated spatial and genomic data, though at a relatively low resolution (spatial data were aggregated at the census tract level and genotype clustering identified using IS6110-based restriction fragment length polymorphism (IS6110-RFLP) analysis)^50^.

Our results are in line with studies of TB in the US^25^ and South Africa^21^ that both noted differences between ‘high-risk’ areas identified with density maps of activity spaces compared to residential locations alone. These studies highlighted the potential importance of activity space analysis, though neither study included genomic data.

Our results are also in line with a recent study in Peru that combined WGS and spatial data to identify differences in activity spaces of genotypically related and unrelated cases and non-TB controls^26^. Notably, this study highlighted the potential to draw on methodology used in spatial ecology, such as using UDs to model activity space, both at the individual and group level^26^. The approach taken in this study was to focus mainly on quantifying size (geographic area) and amount of overlap among participants’ UDs, rather than detecting specific high-risk areas in the community. Our study expanded on these methods by using a spatial point process model, which allowed us to incorporate measures of uncertainty and detect potential hotspots by identifying high-confidence areas of highest spatial intensity.

A limitation of our study is that incorporating activity space may have resulted in including locations that are not relevant to transmission. Another limitation is that we did not include specific measures of temporality, which is also an important element of transmission dynamics.

Another limitation of this study is that we did not examine potential contributing risk factors driving the observed spatial patterns. Spatial variation is often a proxy for the influence of unmeasured variables that may include sociodemographic, social, structural, or environmental factors impacting risk^36,51^. The spatial LGCP modeling approach can incorporate spatially- referenced covariates^51^ (such as population-level sociodemographic characteristics or environmental variables); however these data were not available for our analysis. We focused primarily on identifying geographic areas of increased risk, which could potentially be targeted for outreach such as active case finding. However, further analysis could incorporate additional data or modeling techniques to assess potential risk factors.

Another limitation of this study is an unknown number of missing cases, activity space locations, and WGS data that could potentially alter geographic characterization of genotypic groups.

Although the original study had relatively high enrollment (4,331/5,515 persons diagnosed during the study period), not every person with TB was included, such as those diagnosed but not enrolled and cases that were not detected. In addition, the use of location data obtained through patient interviews is subject to recall bias and underreporting^52^. Other methods of obtaining location data, such as prospective GPS tracking, have been suggested as potential alternatives^26^. However, locations visited during the infectious period prior to diagnosis and study enrollment were of primary interest in this context^53^. Further, a study comparing locations reported by participants and locations captured via GPS loggers found that for three quarters of respondents, over 70% of self-reported locations matched with the GPS data^54^.

## Conclusion

Integrated geospatial and genomic analysis of activity space may help identify potential high-risk locations of sustained transmission in the community. Activity space analysis may improve the geographic characterization of transmission ‘hotspots’ compared to analysis of residential location alone. This could help with planning and mobilizing interventions to interrupt ongoing transmission, and could provide a valuable tool for public health officials working to eliminate TB among marginalized communities^7,10^.

## Data Availability

Data include sensitive identifying information (GPS coordinates for residential locations) and are not made publicly available.

