## Supplementary Figure 1, Supplementary Table 1, Supplementary Figure 2, Supplementary Figure 3, Supplementary Figure 4, Supplementary Figure 5 for "Using genomic epidemiology and geographic activity spaces to investigate tuberculosis outbreaks in Botswana"

**Supplementary Figure 1**. Maximum distance between any two of a participant's own activity spaces, among participants belonging to outbreak groups (≤5 SNP) and ungrouped participants, Gaborone, Botswana, 2012-2016.

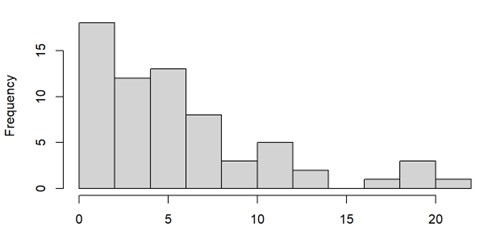

**Supplementary Table 1.** Maximum distance between any two of a participant's own activity spaces, by sociodemographic category among participants belonging to outbreak groups (≤5 SNP) and ungrouped participants, Gaborone, Botswana, 2012-2016.

|  | **Outbreak groups** | **Ungrouped** |
| --- | --- | --- |
| HIV neg | Median 4 (range 1 - 8) | Median 4 (range 1 - 9) |
| HIV pos | Median 4 (range 1 - 10) | Median 4 (range 1 - 9) |
| Income any | Median 4 (range 2 – 10) | Median 4 (range 1 - 9) |
| Income none | Median 3.5 (range 1 – 5) | Median 3 (range 1 - 9) |
| Gender female | Median 4 (range 1 – 6) | Median 4 (range 1 - 9) |
| Gender male | Median 4 (range 1 – 10) | Median 4 (range 1 - 9) |

**Supplementary Figure 2.** Posterior mean estimates of spatial random effect for each outbreak group and subset of 70 randomly selected controls (ungrouped participants) Gaborone, Botswana, 2012-2016. Darker colored areas indicate places where the estimated spatial effect associated with activity spaces for participants in each group was increased after accounting for controls. Values are displayed on the same color scale for all outbreak groups, though on a separate color scale for controls due to difference in sample size.

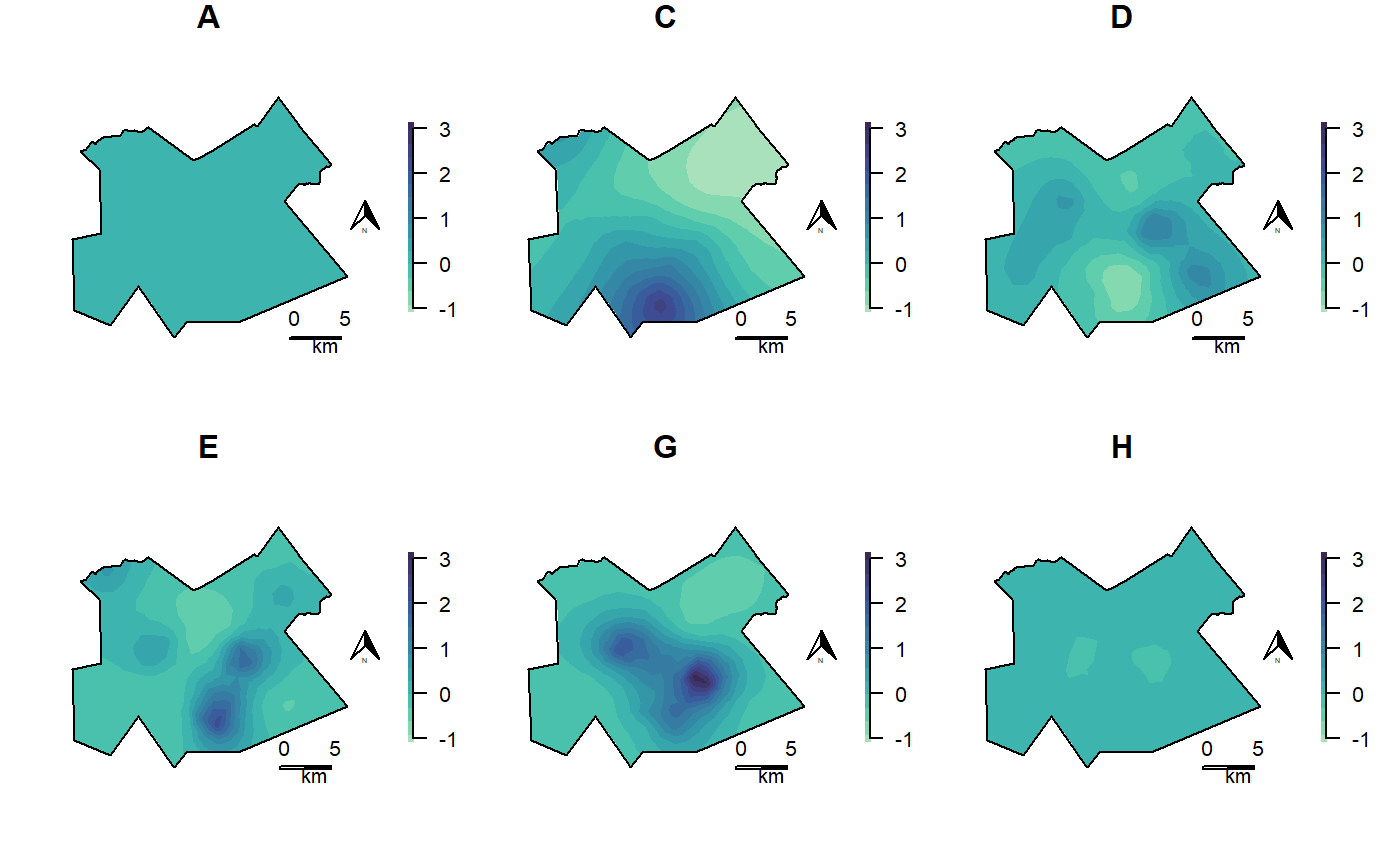

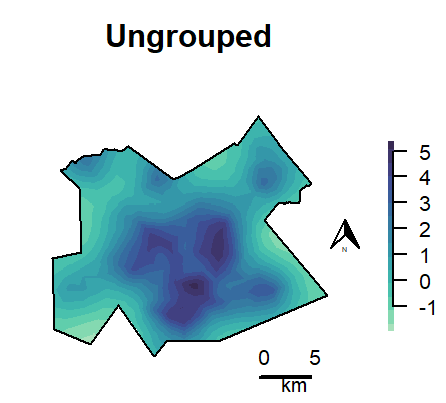

**Supplementary Figure 3.** Predicted mean spatial intensity of activity spaces for participants in each outbreak group and random subset of 70 ungrouped participants, Gaborone, Botswana, 2012-2016. Values are displayed on the same color scale for all outbreak groups, though on a separate color scale for controls due to difference in sample size.

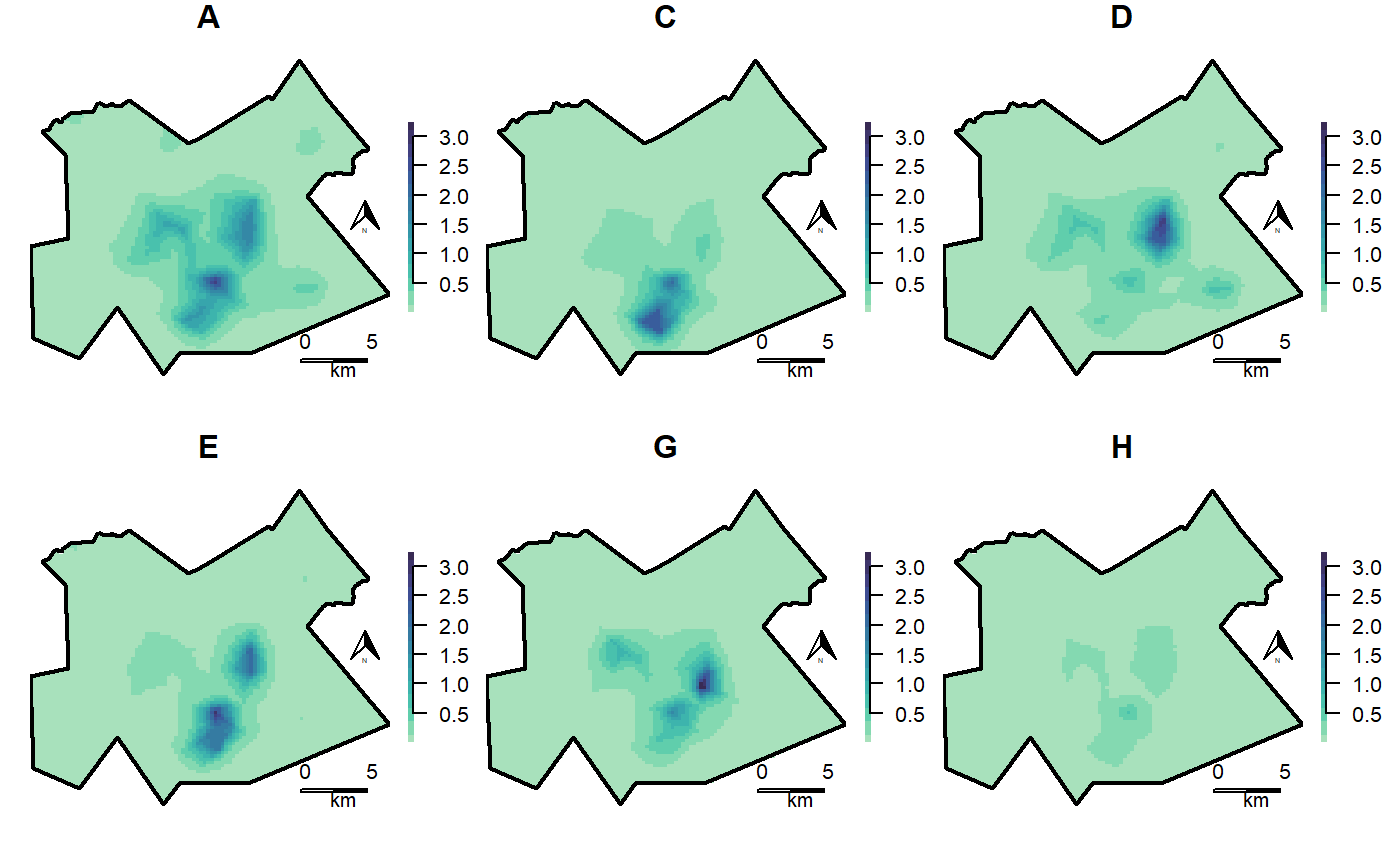

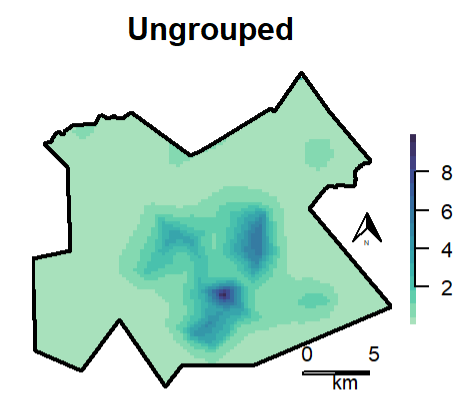

**Supplementary Figure 4.** Posterior mean estimates of spatial random effect for each outbreak group and subset of 140 randomly selected controls (ungrouped participants) Gaborone, Botswana, 2012-2016. Darker colored areas indicate places where the estimated spatial effect associated with activity spaces for participants in each group was increased after accounting for controls. Values are displayed on the same color scale for all outbreak groups, though on a separate color scale for controls due to difference in sample size.

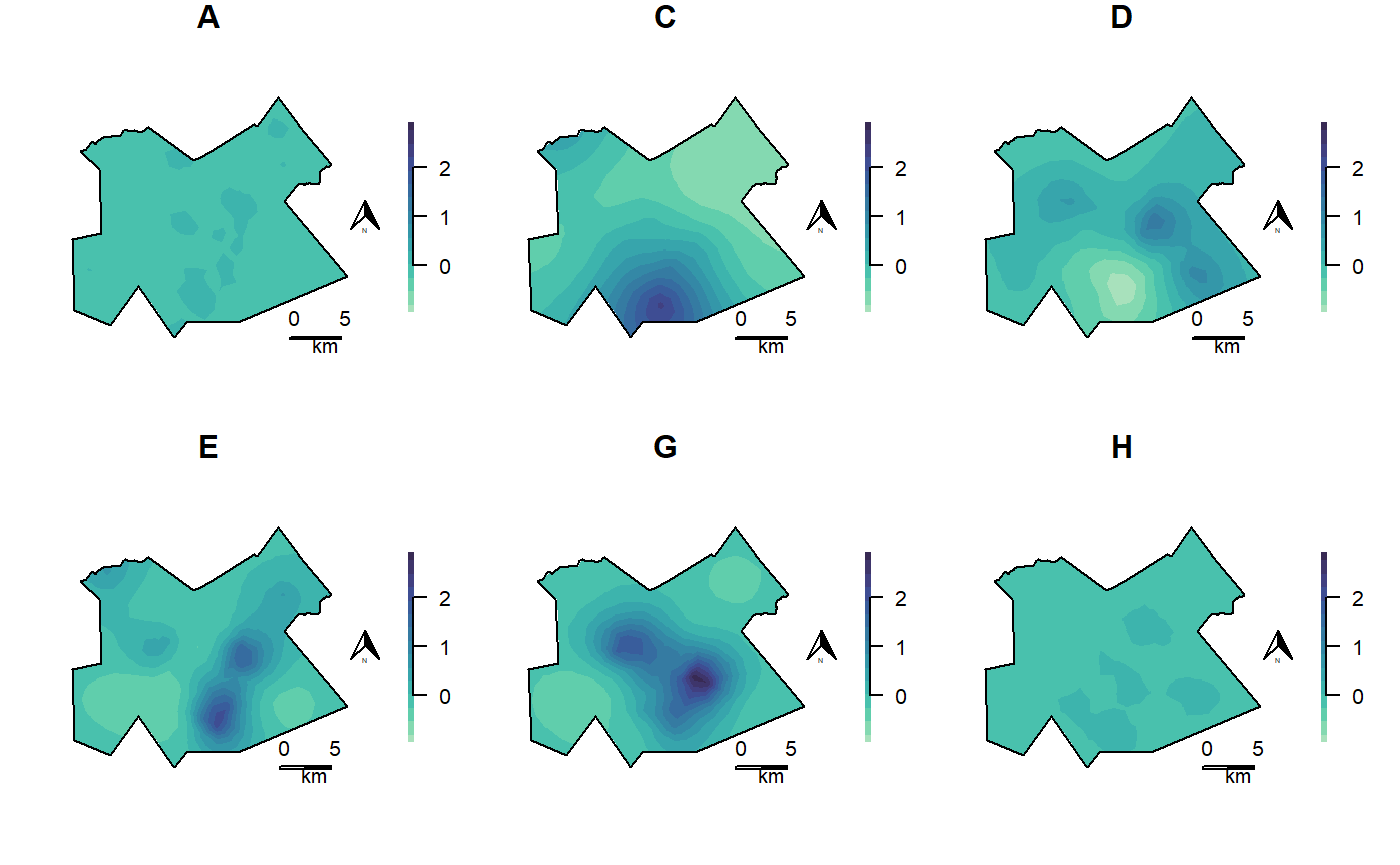

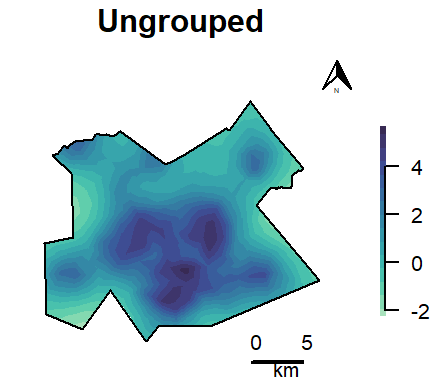

**Supplementary Figure 5.** Predicted mean spatial intensity of activity spaces for participants in each outbreak group and random subset of 140 ungrouped participants, Gaborone, Botswana, 2012-2016. Values are displayed on the same color scale for all outbreak groups, though on a separate color scale for controls due to difference in sample size.

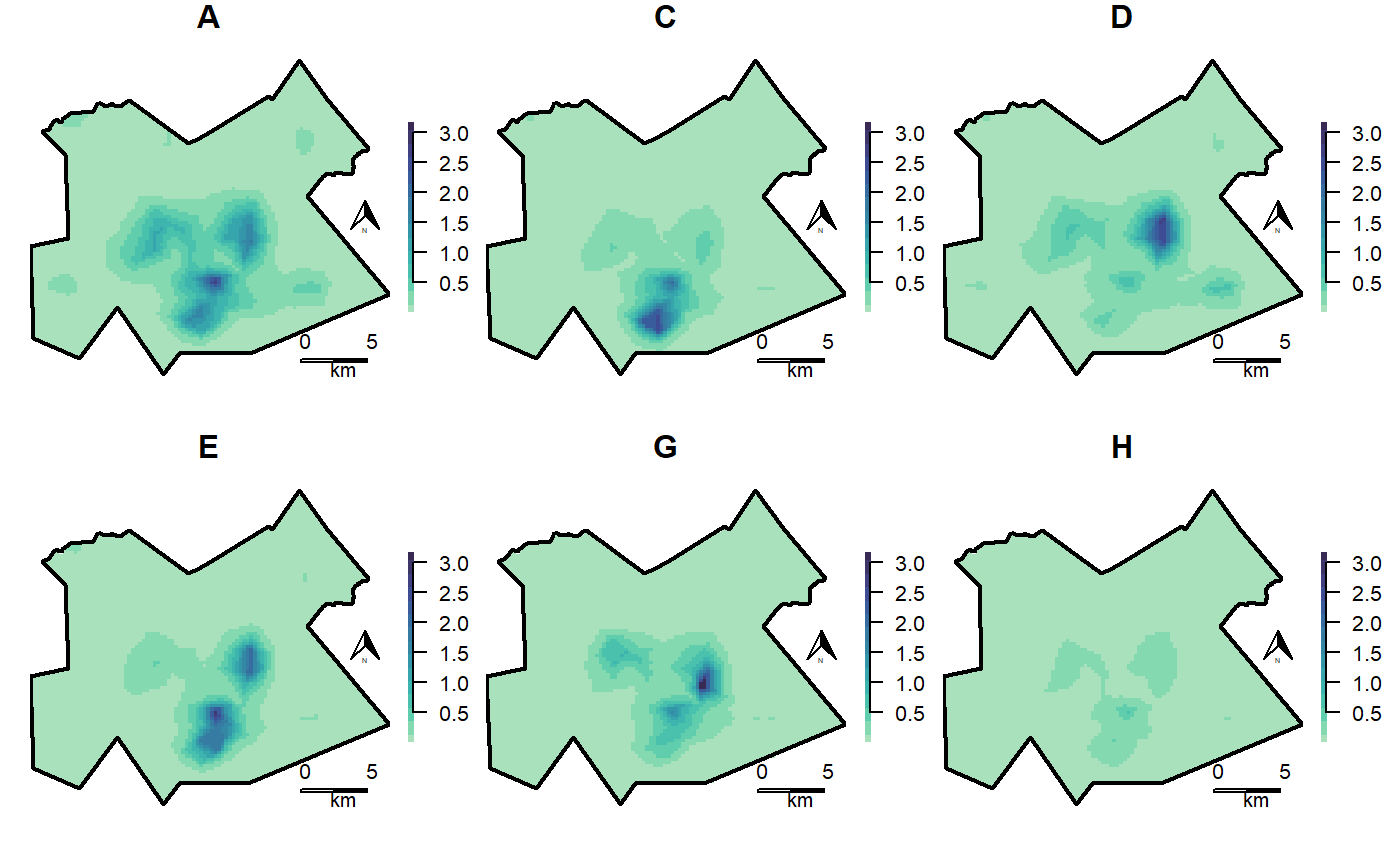

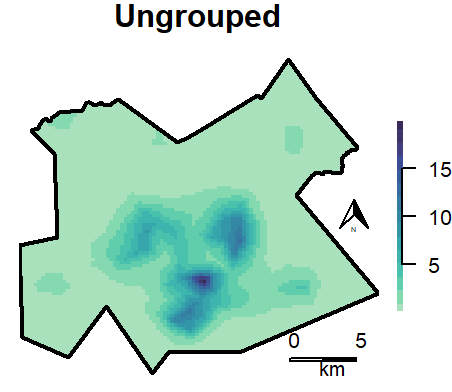
